# India’s Civil Registration System: a potentially viable data source for reliable subnational mortality measurement

**DOI:** 10.1101/2020.04.03.20052894

**Authors:** Chalapati Rao, Mamta Kansal

## Abstract

**Introduction:** The Indian national Civil Registration System (CRS) is the optimal data source for mortality measurement, but is yet under development. As an alternative, data from the Sample Registration System (SRS) which covers less than 1% of the national population is used. This article presents a comparative analysis of mortality measures from the SRS and CRS in 2017, and explores the potential of the CRS to meet these subnational data needs.

**Methods:** Data on population and deaths by age and sex for 2017 from each source were used to compute national and state level life tables. Sex specific ratios of death probabilities in five age categories (0-4, 5-14, 15-29, 30-69, 70 -84, 85+) were used to evaluate CRS data completeness, using SRS probabilities as reference values. The quality of medically certified causes of death was assessed through hospital reporting coverage and proportions of deaths registered with ill-defined causes from each state.

**Results:** The CRS operates through an extensive infrastructure with high reporting coverage, but child deaths are uniformly under reported, as well as female deaths in some states. However, at ages 30 to 69 years, CRS death probabilities are higher than the SRS values in 15 states in males and 10 states in females. SRS death probabilities are of limited precision for measuring mortality trends and differentials. Medical certification of cause of death is affected by low hospital reporting coverage.

**Conclusions:** The Indian CRS is more reliable than the SRS for measuring adult mortality in several states. Targeted initiatives to improve the recording of child and female deaths, to strengthen the quality of medical certification of cause of death, and to promote use of verbal autopsy methods are necessary to establish the CRS as a reliable source of sub national mortality statistics in the near future.

**KEY MESSAGES:**

- The Sample Registration System (SRS) is currently the main source of mortality statistics in India, since the Civil Registration System (CRS) is yet under development
- Limitations in sample size as well as problems with quality of causes of death result in considerable uncertainty in population level mortality estimates from the SRS
- This research evaluated the quality of the sex and age specific mortality risks from the CRS, using the SRS values in each state as reference values
- The CRS has high levels of reporting coverage for death registration, and also measures higher levels of mortality at ages 30 to 69 years in several states, with high precision
- Interventions are required to improve child death registration, strengthen medical certification of cause of death in hospitals, and introduce verbal autopsy for home deaths
- These interventions will establish the CRS as a routine and reliable source for national and subnational mortality measurement in India in the near future

## INTRODUCTION

Mortality measures by age, sex and cause are essential inputs for population health assessment, health policy, and research. Civil registration and vital statistics systems (CRVS) based on medical certification of causes of death are the optimal data source given their legal mandate for total coverage, and design characteristics that potentially assure timeliness and data quality.(1) In particular, the size and dispersion of population in a large country such as India creates the critical need for robust mortality and health measurements at state and even district level. For monitoring progress towards the United Nations Millennium Development Goals (UNSDGs), information on numbers of deaths and death rates from important conditions including tuberculosis, heart diseases, diabetes, cancers, maternal and child health conditions, and road traffic accidents, among others are required each year.(2) These data are required to quantify major health problems, plan and implement health services, monitor their impact, and guide research priorities to reduce deaths and hence improve the health of the population.(3)

The current Civil Registration System (CRS) in India is implemented by the Office of the Registrar General of India (ORGI) housed in the Ministry of Home Affairs under the Registration of Births and Deaths Act, 1969.(4) In anticipation of the long lead time for CRS development as a reliable source for vital statistics, the government of India established a nationally representative Sample Registration System (SRS) in 1970, as an interim population-based data source. In its current form, the SRS operates in 8861 sample units, and covers about 0.5% of the national population, in which births and deaths are continuously recorded by local registrars, with established follow up procedures for quality control and ascertainment of causes of death.(5) Over time, the SRS has served as the optimal data source for estimating mortality indicators for India, principally due to its assurance of high levels of completeness of death registration.(6) Also, since 2001, the SRS uses formal verbal autopsy (VA) methods for ascertaining causes of death.(7) Data from the SRS has been recently used by two different research groups to estimate national and state level cause-specific mortality patterns, using statistical methods to fill gaps arising from limitations with quality of VA recorded causes of death.(8, 9) However, differences in estimation methodology have resulted in considerable variations between the two mortality estimates, and hence limiting their direct utility of either set of estimates for health policy and evaluation. Such data uncertainty underscores the overall limitations of the SRS for cause-specific mortality estimation in India, even at national level.

Under these circumstances, the CRS could serve as a viable source of mortality statistics at national and subnational level, given its universal coverage and other quality assurance elements described elsewhere.(10) In brief, the CRS is based on a comprehensive legal framework, and is implemented in decentralised model through a multi-sectoral network of registration units across urban and rural areas. The system design demonstrates availability of registration infrastructure across the country, uniform operating procedures, established processes for compilation and submission of vital statistics, and routine practices for monitoring performance and data analysis. Annual CRS reports over the past decade indicate a steady improvement in reporting coverage and completeness of death registration.(11)

This article describes findings from a detailed evaluation of the quality of data from the CRS, in terms of reporting coverage, completeness of death registration, and quality of medical certification of cause of death. Where applicable, similar measures from the SRS were used as reference values to evaluate reliability and plausibility of age-sex variations in CRS death registration, both at national and state level for India’s large states. The findings were used to infer the comparative utility of data from the two sources for mortality measurement at sub national level, and to make recommendations for further development of the CRS into a viable source of vital statistics for India.

## DATA SOURCES AND METHODS

The two principal data sources used for this analysis are the annual CRS and SRS vital statistics reports for 2017.(11, 12) The data quality evaluation presents an in depth analysis of reporting coverage, estimated completeness of death registration, and comparative analysis of estimated summary measures of mortality from the two sources, derived from national and state level life tables by age and sex. The evaluation also presents a summary of the information reporting cascade of data from the Medical Certification of Cause of Death (MCCD) Scheme, which is a component of the CRS for registration of deaths attended by physicians.

Reporting coverage of the CRS was extracted from the annual report for 2017, along with data on total population, total numbers of registered deaths, and proportions of delayed registration beyond 1 year from the date of death. The completeness of death registration in the CRS was computed and compared across two methods. The first method, used by the Office of the Registrar General of India (termed as the ORGI method) applied the SRS observed crude death rate in 2017 for each state to the state population estimate for 2017, to estimate the expected deaths that would have occurred in 2017. Completeness was calculated for each state as the proportion of observed CRS 2017 deaths out of these expected deaths.(13) The second completeness estimate was derived from the Adair-Lopez model, which estimates death registration completeness as a function of its relationship with the registered crude death rate, the registered under-five mortality rate, and proportion of individuals aged above 65 years in the study population.(14) The models for these relationships were developed using population and mortality rates from 2451 country years of vital registration data. These input parameters were derived by sex for each state from CRS data for 2017, and used as inputs in the Adair-Lopez model to estimate national and state level completeness by sex.

Life tables for India and all 36 States/UTs were computed from the CRS 2017 data, and for 22 States with larger populations from the SRS data for 2017.(15) The key inputs for life table analyses are population and death distributions by sex and age categories from each data source. For the SRS life tables, these distributions were computed using data from the SRS annual report which contains the following information for each state:

a. total sample population (16)
b. detailed proportionate population distributions by age ((<1 year, 1-4 years, 5-9, 10-14….80-84, 85+) for each sex (17)
c. age-specific death rates according to the same age groups (18)

Subsequently, the state specific SRS population distributions were applied total state population estimates for 2017 to derive age-sex populations as inputs for CRS life tables. Also, the SRS detailed death distributions by age were used to interpolate the coarser age-groups of CRS reported deaths (<1 year, 1-4 years, 5-14, 15-24….55-64, 65-69, 70+) for each state, to derive detailed age-sex numbers of CRS deaths.(19) Detailed age distributions of deaths from Madhya Pradesh were used to interpolate death distributions for Bihar, Jharkhand and Uttar Pradesh, for which the CRS only reported total numbers of deaths for males and females. For the 14 smaller States/UTs, SRS population and death distributions from neighbouring larger states were used to interpolate CRS age-sex distributions of population and deaths. These detailed population and death distributions according to identical categories from each source were then analysed to compute sex specific life tables for India and states for 2017. Summary life table outputs used for comparisons between CRS and SR included life expectancy at birth and probabilities of dying by sex from each source at national/state level across following age groups:

a. birth to 5 years to represent childhood mortality
b. 5 to 14 years to represent school age mortality
c. 15 – 29 years to represent mortality during adolescence and young adulthood
d. 30 to 69 years to represent premature adult mortality
e. 70 to 85 years to represent mortality among the elderly

The variance for each mortality measure from the CRS was calculated according to the Chiang II method, which was incorporated into the spreadsheet calculation tool.(20) The variance of SRS mortality measures was also estimated using the Chiang II method, since cluster-specific death counts were not available for the 8850 sample units of the SRS, hence non-parametric methods could not be applied to estimate variance.(21)

For in-depth understanding the influence of age-specific death registration completeness on the total completeness measures reporting in Table 1 for each state, heat maps were generated to show the ratio of mortality risks between the CRS and SRS by age group for each sex. The SRS measures were used as the reference standard, given its known high levels of completeness of death registration. Ratios were colour coded to reflect lower or higher death recording by the CRS in comparison to the SRS, for the same-age sex group. To understand the impact of precision of mortality estimates from either source on their potential utility for population health assessment and health policy, bar charts were developed to compare risks of premature adult mortality (between 30 and 69 years) along with estimated 95% confidence intervals. Charts were developed separately for males and females, to show mortality risk comparisons between CRS and SRS for states with reported risks that were higher than the national risk estimate from the SRS.

**Table 1:** National and subnational operational characteristics of the Civil Registration and Sample Registration System in India

| Location | Civil Registration system |  |  |  |  |  |  |  | Sample Registration Systems |  |  |
| --- | --- | --- | --- | --- | --- | --- | --- | --- | --- | --- | --- |
|  | Districts | Registration units | Reporting coverage | Population | Observed Deaths | Delayed registration | Estimated completeness of death registration (%) |  | Sample units | Sample population | Observed Deaths |
|  |  |  |  | Millions | 000s | % | AL method * | ORGI method † |  | Millions | 000s |
|  |  |  |  | M / F | M/F | Both sexes | M/F | Both sexes |  | Both sexes | M/F |
| India | 722 | 284671 | 94.2 | 664 / 625 | 3825 / 2628 | 6 | 82 / 76 | 79 | 8850 | 7.925 | 26.6 / 22.2 |
| STATES |  |  |  |  |  |  |  |  |  |  |  |
| 1 Andhra Pradesh | 13 | 14444 | 99.5 | 26 / 26 | 217 / 138 | 3.0 | 93 / 90 | 95 | 325 | 0.327 | 1.3 / 1.01 |
| 2 Arunachal Pradesh | 25 | 221 | 52.9 | 0.69 / 0.64 | 2.2 / 0.74 | 10.2 | 55 / 24 | 36 | 65 | 0.041 | NA |
| 3 Assam | 33 | 676 | 100 | 17 / 16 | 76 / 65 | 11.5 | 72 / 73 | 66 | 300 | 0.219 | 0.76 / 0.59 |
| 4 Bihar | 38 | 9220 | 76.4 | 55 / 50 | 158 / 104 | NA | 56 / 45 | 43 | 330 | 0.366 | 1.05 / 0.99 |
| 5 Chhattisgarh | 27 | 17407 | 100 | 13 / 13 | 97 / 75 | 8.1 | 87 / 88 | 89 | 158 | 0.117 | 0.49 / 0.36 |
| 6 Goa | 2 | 206 | 100 | 1.03 / 1 | 7.8 / 5.2 | 1.1 | 96 / 93 | 103 | 95 | 0.088 | NA |
| 7 Gujarat | 33 | 17572 | 100 | 33 / 31 | 231 / 157 | 1.1 | 87 / 81 | 98 | 478 | 0.426 | 1.52 / 1.14 |
| 8 Haryana | 22 | 608 | 100 | 43784 | 103 / 72 | NA | 92 / 81 | 108 | 256 | 0.241 | 0.82 / 0.56 |
| 9 Himachal Pradesh | 12 | 3285 | N.A. | 3.7 / 3.5 | 23 / 16 | 1.3 | 85 / 83 | 83 | 210 | 0.117 | 0.45 / 0.32 |
| 10 Jammu & Kashmir | 22 | 356 | 63.5 | 6.7 / 5.9 | 22 / 16 | 0.0 | 53 / 59 | 62 | 350 | 0.283 | 0.82 / 0.55 |
| 11 Jharkhand | 24 | 8857 | N.A. | 18 / 17 | 70 / 46 | NA | 72 / 61 | 62 | 211 | 0.157 | 0.39 / 0.46 |
| 12 Karnataka | 30 | 32304 | 100 | 32 / 31 | 292 / 190 | 8.3 | 96 / 94 | 117 | 511 | 0.495 | 1.78 / 1.43 |
| 13 Kerala | 14 | 1035 | N.A. | 17 / 19 | 146 / 117 | 2.0 | 98 / 96 | 108 | 280 | 0.365 | 1.37 / 1.14 |
| 14 Madhya Pradesh | 51 | 24550 | 97.8 | 41 / 38 | 224 / 147 | 7.6 | 69 / 60 | 69 | 448 | 0.402 | 1.52 / 1.16 |
| 15 Maharashtra | 36 | 42015 | 91.2 | 63 / 59 | 369 / 273 | NA | 92 / 90 | 93 | 662 | 0.517 | 1.60 / 1.36 |
| 16 Manipur | 16 | 133 | 95.4 | 1.3 / 1.3 | 2.2 / 1.9 | NA | 34 / 25 | 30 | 165 | 0.149 | NA |
| 17 Meghalaya | 11 | 164 | 62.2 | 1.4 / 1.4 | 8.2 / 5.5 | 3.2 | 84 / 72 | 80 | 130 | 0.081 | NA |
| 18 Mizoram | 8 | 843 | 98.2 | 0.55 / 0.53 | 3.5 / 2.0 | 0.1 | 87 / 70 | 128 | 45 | 0.036 | NA |
|  | Districts | Registration units | Reporting coverage | Population | Observed Deaths | Delayed registration | Estimated completeness of death registration (%) |  | Sample units | Sample population | Observed Deaths |
|  |  |  |  | Millions | 000s | % | AL method * | ORGI method † |  | Millions | 000s |
|  |  |  |  | M / F | M/F | Both sexes | M/F | Both sexes |  | Both sexes | M/F |
| 19 Nagaland | 11 | 1427 | 38.6 | 1.3 / 1.2 | 1.4 / 0.5 | 58.7 | 22 / 14 | 21 | 50 | 0.030 | NA |
| 20 Odisha | 30 | 415 | 100 | 22 / 21 | 184 / 139 | 7.2 | 90 / 90 | 102 | 405 | 0.355 | 1.42 / 1.16 |
| 21 Punjab | 23 | 3202 | 100 | 16 / 14 | 123 / 85 | 1.3 | 95 / 92 | 102 | 249 | 0.206 | 0.83 / 0.60 |
| 22 Rajasthan | 33 | 10085 | 100 | 39 / 36 | 276 / 150 | 16.0 | 88 / 63 | 95 | 350 | 0.321 | 1.11 / 0.81 |
| 23 Sikkim | 4 | 33 | 100 | 0.4 / 0.3 | 2 / 1.4 | NA | 78 / 81 | 113 | 65 | 0.061 | NA |
| 24 Tamil Nadu | 32 | 16458 | 100 | 35 / 35 | 342 / 239 | 3.4 | 97 / 96 | 124 | 545 | 0.518 | 1.95 / 1.50 |
| 25 Telangana | 31 | 8757 | 87.9 | 19 / 19 | 101 / 78 | 1.0 | 82 / 81 | 73 | 224 | 0.209 | 0.74 / 0.62 |
| 26 Tripura | 8 | 1351 | 100 | 2 / 1.9 | 43785 | 16.5 | 93 / 87 | 130 | 90 | 0.106 | NA |
| 27 Uttarakhand | 13 | 8131 | 54.9 | 5.7 / 5.5 | 20 / 14 | 19.1 | 67 / 61 | 47 | 375 | 0.291 | 1.13 / 0.78 |
| 28 Uttar Pradesh | 75 | 56756 | 87 | 117 / 106 | 345 / 227 | NA | 33 / 29 | 38 | 500 | 0.515 | 1.82 / 1.57 |
| 29 West Bengal | 23 | 3945 | 98.8 | 49 / 47 | 256 / 187 | 4.9 | 85 / 82 | 92 | 555 | 0.544 | 1.68 / 1.35 |
| UNION TERRITORIES |  |  |  |  |  |  |  |  |  |  |  |
| 1 Andaman & Nicobar | 3 | 91 | 100 | 0.30 / 0.26 | 1.3 / 0.8 | 3.2 | 60 / 51 | 74 | 55 | 0.046 | NA |
| 2 Chandigarh | 1 | 13 | 100 | 1.0 / 0.85 | 13.5 / 7.7 | 0.1 | 98 / 97 | NA | 40 | 0.032 | NA |
| 3 Dadra & Nagar Haveli | 1 | 23 | 100 | 0.25 / 0.29 | 1.3 / 0.82 | 1.4 | 72 / 70 | 107 | 35 | 0.041 | NA |
| 4 Daman & Diu | 2 | 24 | 100 | 0.22 / 0.14 | 0.73 / 0.41 | 2.3 | 49 / 51 | 69 | 25 | 0.028 | NA |
| 5 Delhi | 11 | 18 | 100 | 11.8 / 10.3 | 84 / 52 | 0.9 | 91 / 85 | NA | 199 | 0.133 | 0.30 / 0.21 |
| 6 Lakshadweep | 1 | 10 | 100 | 0.042 / 0.040 | 351 | 0.3 | NA | 65 | 14 | 0.013 | NA |
| 7 Puducherry | 4 | 36 | 100 | 0.9 / 0.9 | 8.3 / 5.1 | 0.4 | 97 / 93 | 104 | 55 | 0.050 | NA |

\* Adair - Lopez method (ref)
† Office of Registrar General of India method, uUsing SRS crude death rate to calculate expected deaths

Finally, to evaluate the quality of data on causes of death from the CRS, a descriptive table of the performance of the Medical Certification of Cause of Death Scheme was developed. The table shows for each state the cascade of cause of death reporting under this scheme, starting with the number of accredited hospitals, the number of hospitals covered by the MCCD regulations, the institutions that submitted annual reports in 2017, the eventual proportions of registered deaths in each state that were medically certified as to cause, and the proportions of these certified deaths that were assigned non-specific conditions as underlying causes of death. The table also reports an estimate of the proportion of SRS deaths recorded in 2017 that received some form of medical attention at death, from either a government or private hospital.(22) This statistic indicates the proportion of deaths for which there is a likelihood of obtaining some medical inputs into the likely condition(s) that could have caused the death.

## RESULTS

### Reporting coverage and completeness

A broad overview of the operational characteristics of the Civil Registration and Sample Registration Systems is presented in Table 1, as a basis for understanding subsequent analyses. The CRS is implemented through an extensive network of registration units across the country, with high levels of reporting coverage in almost all states. The estimates of death registration completeness correlate well across the two methods for all states, and with coverage levels, except in Madhya Pradesh and Uttar Pradesh which show low completeness despite high reporting coverage. This indicates that a number of registration units in these states had submitted reports with considerably less than expected death registrations. A more detailed analysis across districts in these states could help identify locations where interventions could be required to improve CRS performance. Delayed registration is generally low, but data is not available for some states, and this could influence the validity of annual mortality measures reported from the CRS.

The SRS does not experience problems with reporting coverage or completeness of death registration, given its strengths in design and performance, as reported elsewhere.(6) However, a key drawback is the limitations from sample size. While there are large numbers of sample sites in each state, the low population coverage and resultant low numbers of observed deaths is indicative of the relatively small size of each cluster, which could limit the precision of mortality indicators by sex, age and location, as compared to the CRS. For states with smaller populations, the SRS does not even report schedules of observed age-specific death rates probably o account of these sample size limitations, which precludes the computation of life tables and their outputs.

### Summary mortality measures

Table 2 shows summary mortality measures from the two sources at national and state level. We first compared estimated life expectancies at birth, and found that CRS life expectancies for males and females are less than or equal to those from the SRS in Karnataka, Kerala and Tamil Nadu. Given that all these states have very high levels of CRS death registration completeness, this indicates that the SRS is potentially under estimating mortality in these states. The CRS life expectancies for males are within a 1 year margin of the SRS levels including Andhra Pradesh, Gujarat, Haryana, Maharashtra, Odisha, Punjab and Rajasthan. However, CRS female life expectancies in these states are considerably higher that SRS levels, suggestive of under reporting of female deaths in the CRS.

**Table 2:**
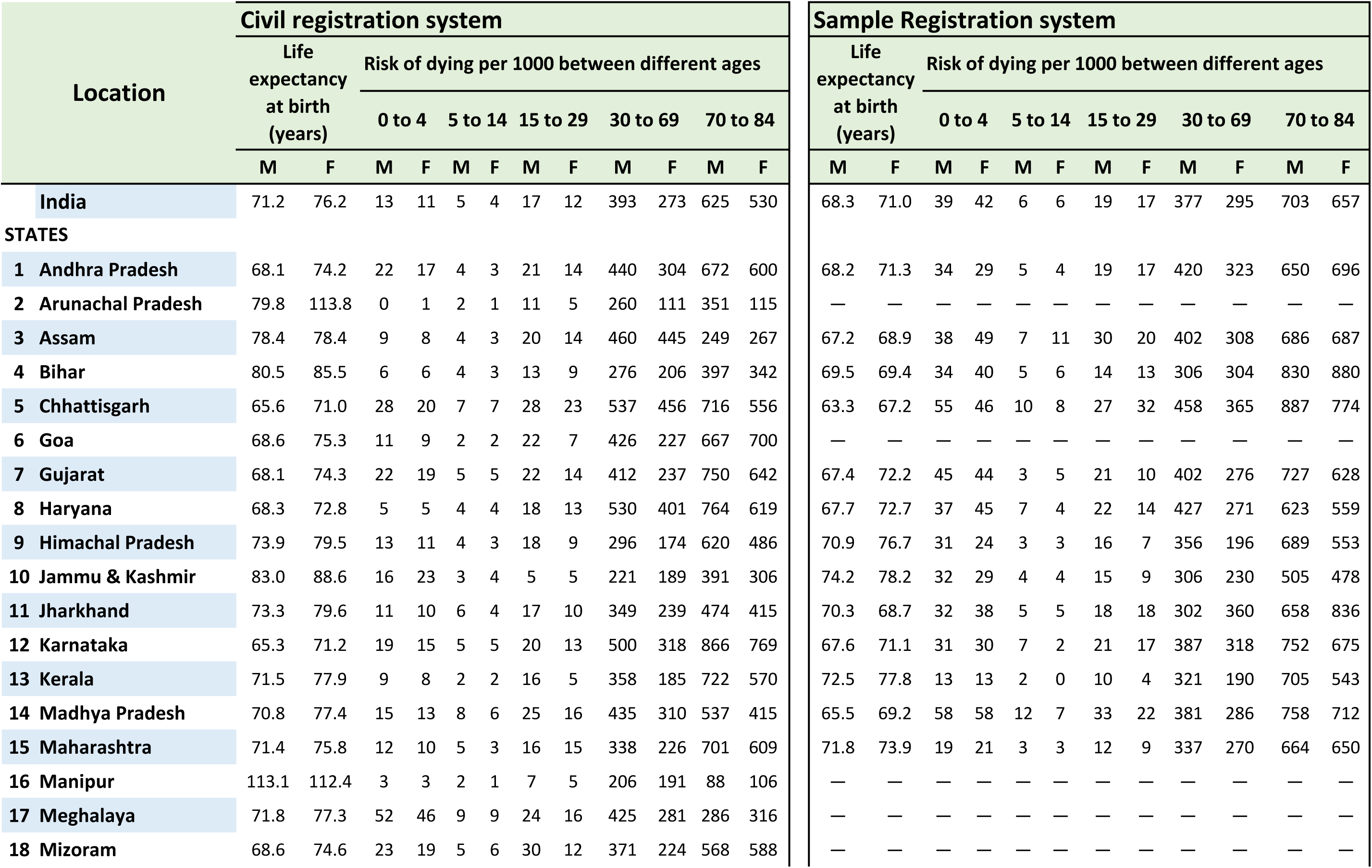
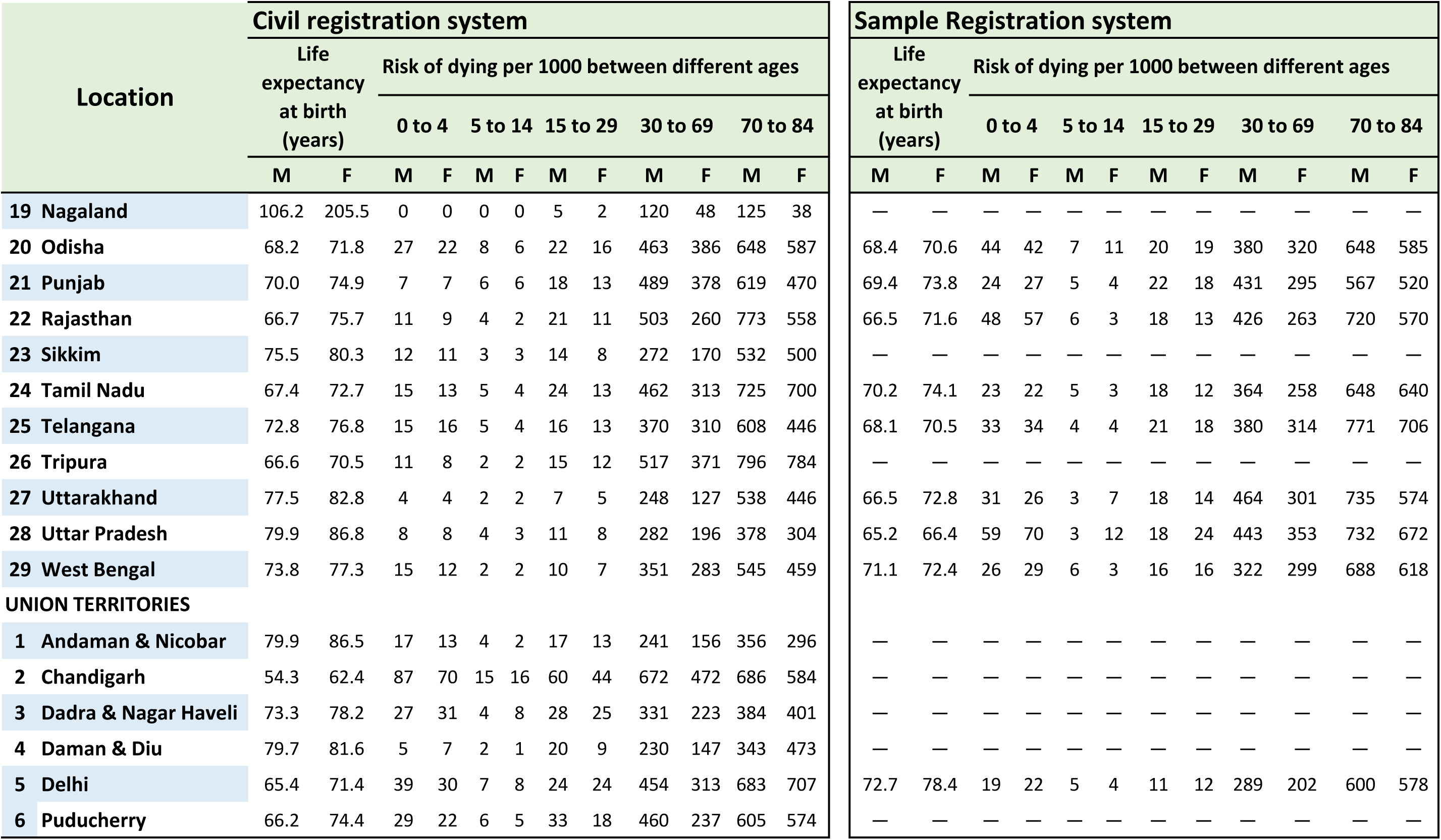
National and subnational summary measures of mortality for 2017 fom the Civil Registration and Sample Registration Systems in India

All other states with data from both sources show higher CRS life expectancies for both males and females, which indicate lower death recording in the CRS as compared to that in SRS sites in these states. Further, the CRS life expectancy estimates are implausibly high (>75 years in males, and >80 years in females) for the states where comparison measures from the SRS are not available, including Arunachal Pradesh, Sikkim, Manipur and Nagaland, and the Union Territories of Andaman and Nicobar Islands, and Daman and Diu. On the other hand, the lower life expectancies and high age-specific mortality risks observed in the Union Territories of Chandigarh, Delhi and Puducherry are a reflection of the practice of death registration at place of occurrence. The CRS data for these locations include deaths that occur in individuals from neighbouring states who seek terminal medical care in the advanced tertiary hospitals in these cities, hence inflating the numerators for calculating mortality indicators. Life tables could not be computed for Lakshadweep owing to low numbers of population and registered deaths by age and sex.

Table 2 also shows the estimated probabilities of dying by sex and age from the two data sources. As mentioned previously, the SRS death probabilities by sex and age are routinely used for mortality estimation for India and States, given their reputed reliability. In summary, they demonstrate considerable inter-state variations in mortality patterns. For instance, there is a five-fold difference in risk of child mortality, ranging from 13 per 1000 in Kerala to about 65 per 1000 risk in Uttar Pradesh, for male and female children together. There is similar variation in adult mortality risks across states along with considerable gender differentials, ranging between 289 and 464 per 1000 population in males, and between 190 and 365 per 1000 in females.

We used the SRS death probabilities as reference values to compute the ratios of completeness of CRS sex and age specific death probabilities (see Figures 1 and 2). Ratios less than 1 are indicative of under-reporting by the CRS, and ratios above 1 indicative higher mortality recording by the CRS. As can be seen, there is general under-reporting of childhood mortality in the CRS across all states, with ratios less than 0.5 in about three-fourths of states. On the other hand, the probability ratios show higher level of CRS death recording at adult ages (30 to 69 years) in most states, but particularly so for males. Even the states of Assam, Madhya Pradesh and Telangana which very low levels of overall completeness (<80%) show high death probability ratios at ages 30 to 69 years, in both males and females. Similarly, the states of Chhattisgarh, Jharkhand and West Bengal have low completeness levels (80-90%), but show ratios of more than 1 at these ages, but for males only. Several states also demonstrate higher CRS death recording among the elderly. In general, the female death probability ratios are less than 1 in categories for ages above 15 years in many of the states, indicative of a general under reporting of adult female deaths in the CRS.

**Figure 1:**
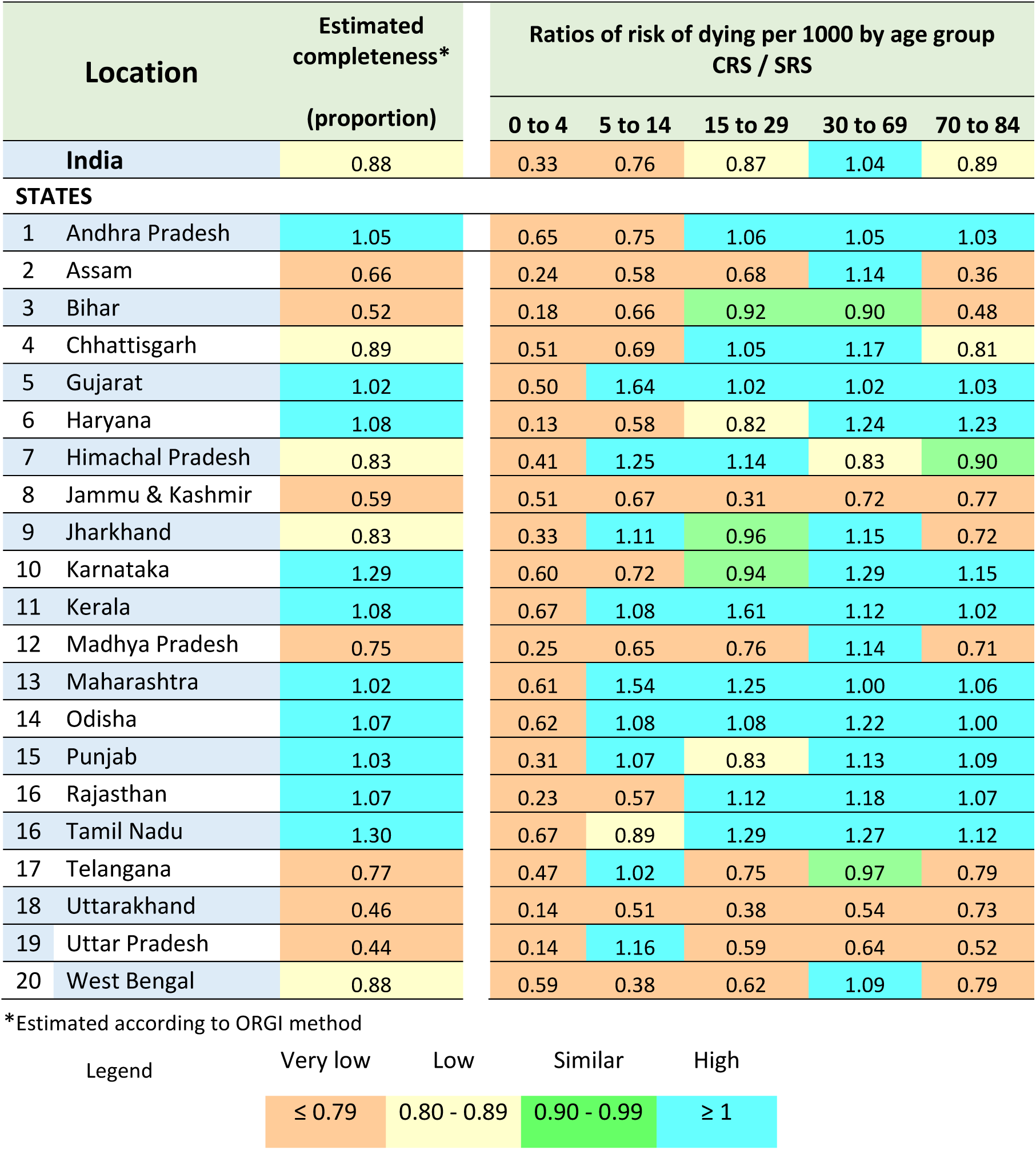
Ratios of age-category probabilities of death in males from CRS and SRS by state in 2017.

**Figure 2:**
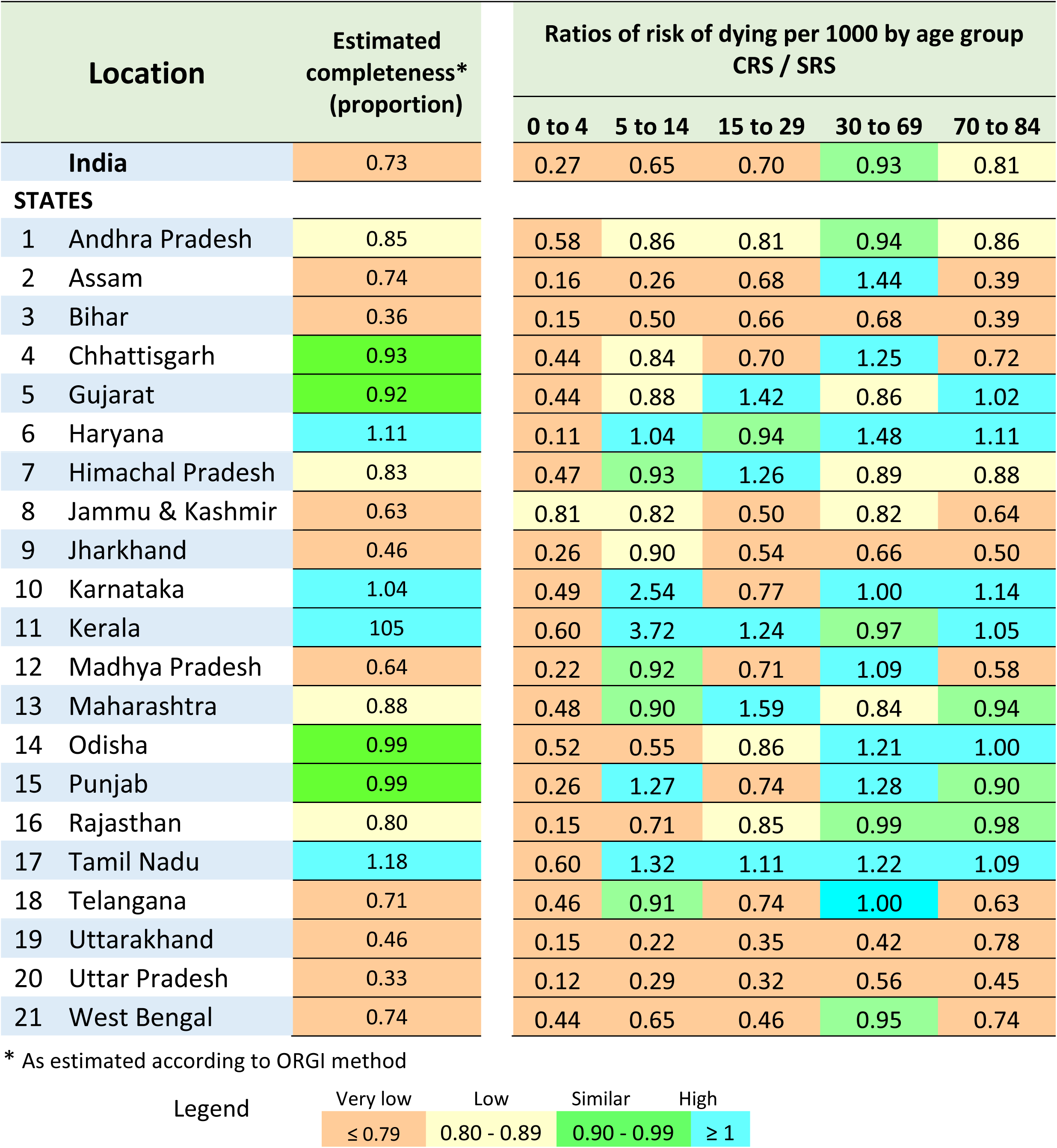
Ratios of female age-category probabilities of death from CRS and SRS by state in 2017.

Figures 3 and 4 show the differentials between CRS and SRS death probabilities at ages 30 to 69 years, for states with CRS measures that are higher than the SRS. The 95% confidence intervals also display the influence of limited size of the SRS sample, in terms of the low precision of the estimates. The graphs clearly suggest that the SRS appears to be grossly under-estimating adult mortality risks in many states in both sexes. For males, 15 states show higher CRS death probabilities, with statistically significant differences in 10 of these states. In females, the differences are statistically significant in 6 out of 10 states with higher CRS death probabilities. As mentioned before, the SRS variance measures do not account for cluster specific death count variations, since these data were not available. Measurement of this additional error would further widen the 95% confidence intervals for the SRS estimates, and accentuate the already low levels of precision in SRS age-specific mortality risks. This would further limit the utility of the SRS estimates for understanding levels, trends and differentials across the states. On the other hand, the very narrow 95% confidence intervals for CRS mortality risks for all states suggest that the CRS sample sizes are likely to be adequate for reasonably precise measurement of these indicators even at district level.

**Figure 3:**
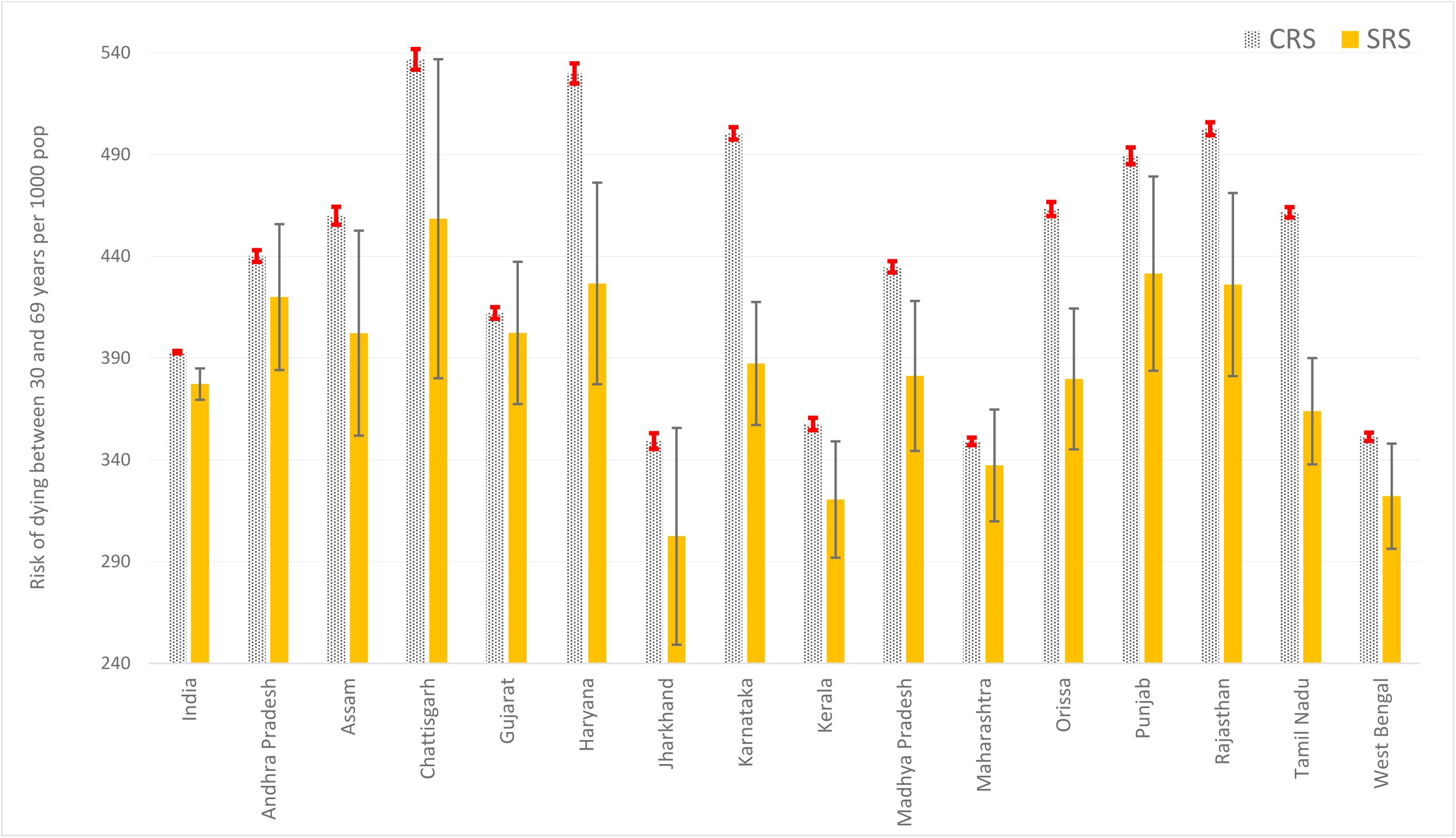
Comparisons of risk of dying between 30 and 69 years from CRS and SRS for males in India and selected states, 2017.

**Figure 4:**
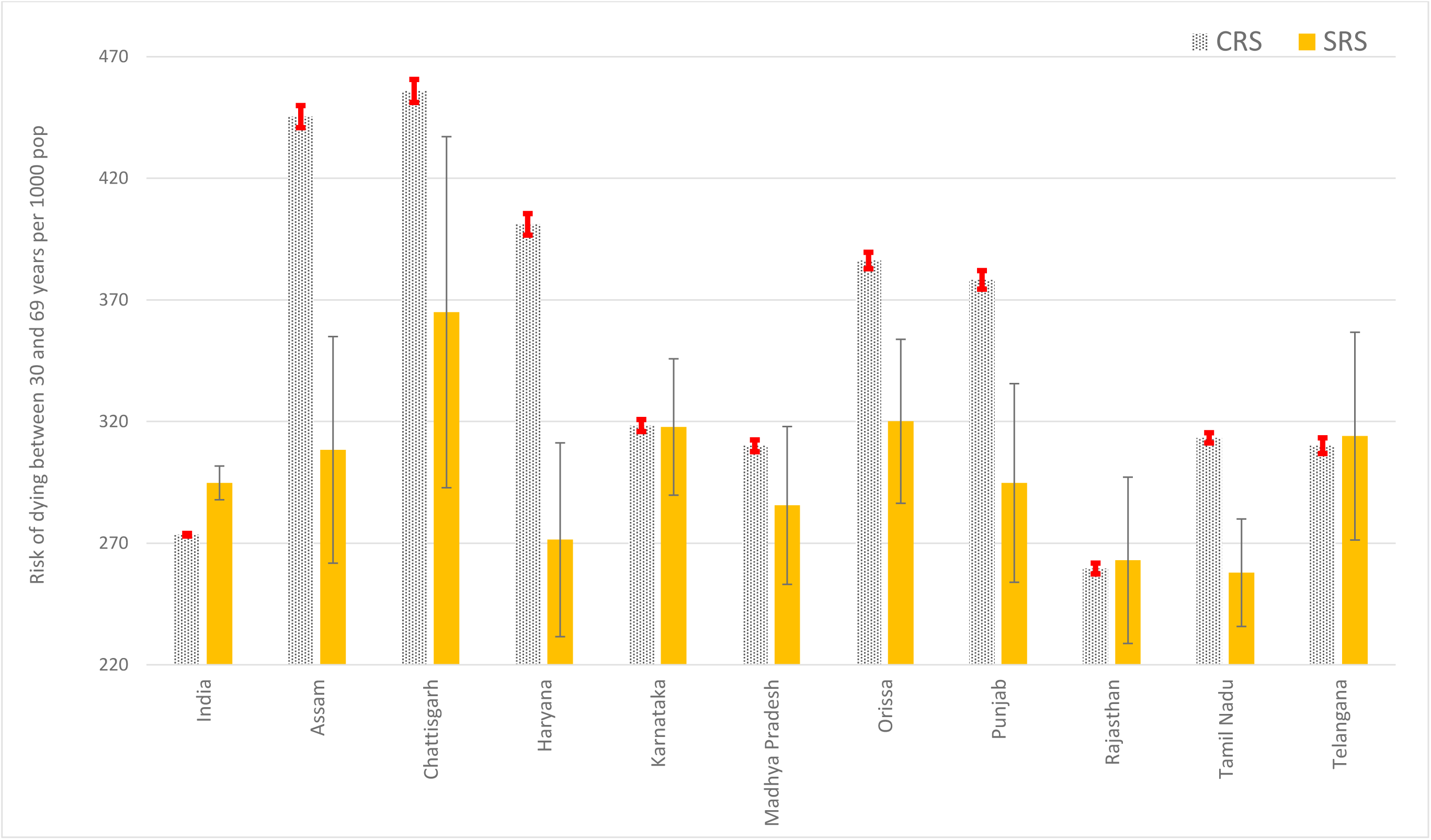
Comparisons of risk of dying between 30 and 69 years from CRS and SRS for females in India and selected states, 2017.

### Medical certification of cause of death

An assessment of the availability and quality of reported causes of death provides an epidemiological perspective of the utility of data from the CRS. Table 3 shows the information cascade under the Medical Certification of Cause of Death Scheme (MCCD) for 2017. It can be readily perceived that there is considerable heterogeneity in reporting performance across states. At one level, only 71% of registered hospitals across the country are covered under the MCCD Scheme, with markedly low proportions in Bihar, Kerala, Punjab, Uttarakhand, and West Bengal. Among larger states, this variable was not reported for Jammu and Kashmir, Jharkhand, Madhya Pradesh and Uttar Pradesh. At the next level, there is equal concern arising from the widespread shortfall in submission of MCCD data by the covered hospitals across all states in 2017. Among the larger states, only Assam, Goa, Maharashtra and Rajasthan achieved a 100% institutional reporting coverage. Very low institutional reporting (<50%) was observed from Andhra Pradesh, Gujarat, Haryana, Himachal Pradesh, Telangana and West Bengal, and this variable could not be computed for several other large states notably Madhya Pradesh and Uttar Pradesh, because of missing data. The net effect of these shortfalls in administrative and reporting coverage results in only 22% of registered national deaths in 2017 being reported with a medically certified cause of death, with a range from 5% in Jharkhand to 100% in Goa. Another parameter for assessing data quality is the percentage of deaths certified with only symptoms or non-specific terms such as senility as causes of death. This was found to be at or above the international threshold of 10% in 14 out of 36 States/Union Territories, ranging from 1 to 43 % across the states.(23)

**Table 3:** Reporting performance of the Medical Certification of Cause of Death Scheme, 2017

| Location | Total registered deaths | Medical attention at death* | Total registered Hospitals | Hospitals covered by MCCD | Hospitals reporting MCCD | Proportion of MCCD deaths | MCCDs with ill-defined causes |
| --- | --- | --- | --- | --- | --- | --- | --- |
|  | Number | % | Number | Number | Number | % | % |
| **India** | 6,462,977 | 44 | 82,452 | 58,240 | 41,404 | 22 | 10 |
| **STATES** |  |  |  |  |  |  |  |
| 1 Andhra Pradesh | 355,546 | 46 | 2,483 | 2,483 | 965 | 14 | 10 |
| 2 Arunachal Pradesh | 2,137 | NA | 188 | 188 | 41 | 42 | 43 |
| 3 Assam | 141,012 | 35 | 1,209 | 1,209 | 1,209 | 31 | 1 |
| 4 Bihar | 261,425 | 31 | 683 | 42 | 42 | 7 | 10 |
| 5 Chhattisgarh | 175,035 | 40 | 905 | 693 | 400 | 19 | 18 |
| 6 Goa | 13,020 | NA | 174 | 174 | 174 | 100 | 9 |
| 7 Gujarat | 388,316 | 44 | 1,542 | 1,542 | 67 | 21 | 30 |
| 8 Haryana | 174,937 | 37 | 3,329 | 1,162 | 452 | 16 | 17 |
| 9 Himachal Pradesh | 39,114 | 66 | 438 | 438 | 120 | 15 | 12 |
| 10 Jammu & Kashmir | 37,184 | 55 | 592 | NA | NA | NA | NA |
| 11 Jharkhand | 116,393 | 33 | 920 | NA | NA | 5 | 1 |
| 12 Karnataka | 481,747 | 50 | 4,345 | ... | 1,719 | 30 | 2 |
| 13 Kerala | 263,342 | 79 | 2,519 | 150 | 150 | 11 | 3 |
| 14 Madhya Pradesh | 370,538 | 55 | 5,685 | NA | NA | 9 | 7 |
| 15 Maharashtra | 647,161 | 49 | 25,011 | 25,011 | 25,011 | 39 | 12 |
| 16 Manipur | 4,130 | NA | 119 | 38 | 16 | 55 | 13 |
| 17 Meghalaya | 13,779 | NA | 164 | 170 | 170 | 41 | 7 |
| 18 Mizoram | 5,525 | NA | 103 | 103 | 100 | 55 | 5 |
| 19 Nagaland | 1,836 | NA | 11 | 11 | 5 | 15 | 0 |
| 20 Odisha | 322,660 | 47 | 741 | 659 | 490 | 12 | 9 |
| 21 Punjab | 210,398 | 56 | 1,515 | 472 | 264 | 16 | 7 |
| 22 Rajasthan | 424,763 | 48 | 2,963 | 2,963 | 2,963 | 13 | 4 |
| 23 Sikkim | 3,339 | NA | 32 | 32 | 32 | 44 | 3 |
| 24 Tamil Nadu | 580,496 | 46 | 7,405 | 6,841 | 5,475 | 43 | 6 |
| 25 Telangana | 178,345 | 44 | 7,827 | 7,144 | 284 | 33 | 7 |
| 26 Tripura | 26,296 | NA | 124 | 124 | 124 | 21 | 11 |
| 27 Uttarakhand | 33,931 | 54 | 1,216 | 36 | 23 | 7 | 6 |
| 28 Uttar Pradesh | 571,170 | 49 | 9,503 | NA | NA | 9 | 6 |
| 29 West Bengal | 442,995 | 45 | 463 | 153 | 56 | 13 | 12 |
| **UNION TERRITORIES** |  |  |  |  |  |  |  |
| 1 Andaman & Nicobar | 2,137 | NA | 37 | 37 | 37 | 62 | 3 |
| 2 Chandigarh | 21,236 | NA | 35 | 13 | 5 | 44 | 7 |
| 3 Dadra & Nagar Haveli | 2,080 | NA | 33 | 11 | 11 | 58 | 7 |
| 4 Daman & Diu | 1,136 | NA | 13 | 13 | 13 | 87 | 1 |
| 5 Delhi | 136,117 | 66 | NA | NA | 900 | 61 | 24 |
| 6 Lakshadweep | 351 | NA | 13 | 13 | 13 | 95 | 13 |
| 7 Puducherry | 13,350 | NA | 112 | 73 | 73 | 64 | 23 |

\* Data from the SRS 2017 report

Finally, to envisage the potential reach of the MCCD Scheme, Table 3 also reports the estimated proportion of deaths that had received medical attention in a government or private hospital immediately before death for each state. These estimates were reported by the SRS for 2017, which captures this information for every death recorded by the system. The national average for this variable is 44%, with most states reporting values closely clustered around the national average. This suggests that a well-functioning MCCD Scheme could readily double the proportion of nationally registered deaths with medically certified causes, which would considerably improve the overall utility of cause-specific mortality data from the CRS.

## DISCUSSION

Reliable measurement of cause-specific mortality is among the first principles of evidence-based population health policy, along with descriptive analyses of major health problems, and assessments of facilities and human resources available for health service delivery. For India, there is an imperative need for reliable mortality data to measure key UNSDG and related international health indicators at state level, and in some cases, even at district level. The analyses presented here establish several important facts about the status of current mortality statistics programs in India, and provide a scientific basis for specific interventions to improve data quality to meet these needs.

An important preliminary finding is that the SRS is clearly inadequate for measuring sex-age specific mortality indicators at state level in India. In particular, the SRS under estimates mortality risks for ages between 30 to 69 years in several states, and this is a cause of concern, given the need for accurate mortality measurement in this age group to address the burden from non-communicable diseases (24) This is coupled with the poor levels of precision of these indicators from the SRS as seen in Figure 3 and 4, arising from grossly insufficient sample size, which precludes the reliable assessment of mortality differentials across states or over time. On the other hand, the Civil Registration System provides a distinct advantage in terms of precision, owing to the enhanced effect of total population coverage of the system, which eliminates sampling error. Moreover, the very high levels of reporting coverage coupled with high estimated completeness for most states as reported in Table 2 effectively support an inference of better precision and reliability of CRS measures of adult mortality. Even for states with lower levels of estimated completeness, the large numbers of registered deaths suggest the potential for some districts within these states to have better levels of completeness (>85%), which would enable more reliable estimation of adult mortality for these districts. From a health policy perspective, the higher than expected levels of adult mortality observed from CRS data indicate a greater need for health systems strengthening for programs to control non-communicable disease and other major causes of adult deaths in India.(25)

At the same time, it should also be noted that the CRS data could be biased on account of delayed registration, arising from deaths that actually occurred in previous years being registered and counted as deaths in 2017. This information is not available for all states (see Table 1) and hence could not be estimated at national level. However, these proportions are relatively low for most states which provided these data, and similar trends in delayed registration are observed from previous annual CRS reports during 2014-2016, resulting in a similar carry-over of deaths from year to year. Hence, we believe that these factors minimise the potential for bias from delayed registration on CRS mortality measures for 2017.

From an administrative standpoint, Table 2 demonstrates the huge scope and magnitude of CRS operations in India, as inferred from the availability of infrastructure deployed across 284,671 primary registration units, which implement standardised operations for death registration, despite the heterogeneity of structure and design at state and local level.(10) This is combined with apparently efficient processes for management and compilation of registration data in most states, resulting in timely reporting and dissemination of vital statistics within 18-20 months of the reference period. Even through the estimated coverage of the MCCD Scheme is low, annually over 1.4 million deaths are registered with a medically certified cause. Reporting of MCCD data from all states is indicative of the presence of local capacity across the country, and hence potential for decentralised scaling up of system strengthening interventions.

Despite these structural advantages, there are several key limitations of both registration and statistical operations in the CRS. For instance, it is clearly evident that there is under-reporting of child deaths. One potential reason could be the length of the reporting period for birth registration, which is 21 days. Since the majority of infant deaths occur within the first three weeks, this limits the likelihood of bereaved families to report both the birth and death. For this reason, an alternate reporting model could be developed, through involvement of health sector institutions and staff in active notification of infant deaths. There is also a need to conduct social science research to investigate factors for the apparently lower levels of female death registration in most states. Next, mortality measures are not reliable for states with reporting coverage and/or estimated completeness levels which are less than 80%. These states would require a detailed assessment of the design, resources, management and operations of the CRS at state level, according to a standard international framework for such assessments.(26) Where feasible, a thorough evaluation of district level mortality indicators should be undertaken according to the methodology reported here, to identify nodes that require specific attention through system strengthening interventions.

A third key limitation lies in the performance of the MCCD Scheme, ranging from low reporting coverage of hospitals to low quality of reported causes of death. To address reporting coverage, the Registrar-General of India has issued instructions in 2014 to extend the MCCD Scheme to cover all government, private, and not-for-profit medical institutions across the country, including a special note to maintain updated lists of such medical institutions in each state.(27) A targeted initiative is required to implement these instructions and close this performance gap, through close monitoring of MCCD reporting in all states, supported by teaching programs to strengthen quality of cause of death certification.(28) To improve the overall quality of cause attribution at death registration, there is also an urgent need to implement verbal autopsy methods for home deaths without medical attention. As demonstrated by the SRS, as well as its precedence in the erstwhile ORGI Survey of Cause of Death Scheme in Rural areas (SCD-Rural) which operated during 1965-1999, there is considerable experience in using VA methods across India.(4, 7) In particular, the SCD-Rural Scheme demonstrated a viable working collaboration between the administration and health sectors at the level of the Primary Health Centre (PHC). Each PHC serves an average of 25,000 population, with about 150-200 deaths each year. Considering that about 20% of these deaths would have a medically certified cause, this would leave the PHC staff with a VA workload of about 2-3 cases per week which appears logistically feasible, when shared across sub centres. This PHC based model for rural areas could be combined with a strategy for using VA for deaths occurring without medical attention in urban areas.

From a broader perspective, these VA activities should be embedded within a broader CRS strengthening initiative which also includes activities to improve registration completeness, medical certification, and data management and analysis for local empirical mortality measurement, starting with representative samples of districts from each state.(29, 30) Such an initiative would be well within the scope of the recent ORGI instructions to strengthen infant death recording, closely monitor registration levels and calculate vital rates at state and district level commencing from 2018. These instructions include a justification that CRS data are ‘exact and real data certified by registering authorities and are therefore legally admissible’ for such calculations.(31)

## CONCLUSIONS

In conclusion, the analyses presented in this article clearly demonstrate that India’s Civil Registration System has major strengths in infrastructure, reporting coverage, data completeness and management. In several states, these strengths also translate into more reliable CRS based adult mortality risks than those from the SRS, hence establishing reliable mortality baseline measures for monitoring non-communicable disease mortality at state and district level in India. Looking forward, there appears to be sufficient political will and support for further reforms to strengthen local registration and statistical operations as well as medical certification of causes of death, in the form of recent government regulations on these subjects. All these observations indicate that with appropriately designed system strengthening initiatives, the Indian CRS could be able to serve its goal as a reliable source for national and sub national vital statistics in the near future.

## Data Availability

All data are available in the public domain with the sources listed among the references

